# Patterns of Rising HIV Positivity in Northern Madagascar: Evidence of an Urgent Public Health Concern

**DOI:** 10.1101/2023.07.02.23291171

**Authors:** Kyle E. Robinson, Jackson K. Long, Mamantsara Fardine, Adriantiana M. Stephano, Eric P. Grewal

## Abstract

Despite over two decades of progress against HIV/AIDS in adjacent sub-Saharan Africa, HIV rates and deaths due to AIDS are exponentially rising in Madagascar. Furthermore, a growing body of evidence suggests that, due to a scarcity of general-population screening data, even the startling increase demonstrated by official models vastly underestimates the true population prevalence of HIV. Thus, we aimed to implement a real-world HIV screening and treatment protocol to serve a general population stemming from across Northern Madagascar. In collaboration with the Malagasy Ministry of Health, we provided point-of-care HIV screening and confirmatory testing for over 1,000 participants from 73 towns, villages, and cities recording an overall HIV prevalence of **2.94%**. Notably, we observed a **13.1%** HIV prevalence rate among urban populations and showed that proximity to a major route of travel was significantly associated with HIV risk. We also observed a link between HIV risk and various occupations, including those associated with increased mobility (such as mining). Importantly, all HIV-positive individuals were initiated on antiretroviral therapy in concordance with local health authorities. To our knowledge, this study marks the largest primary test data-based HIV study to date among Madagascar’s general population, showing a greatly higher HIV prevalence (3.0%) than previously reported modeling based figures (0.3%). Our rates aligned with the pattern of higher prevalence demonstrated in smaller general-population screening studies occuring more commonly prior to political strife in the mid-2000s. These findings demonstrate evidence of a growing HIV epidemic in Northern Madagascar and underscore the need for future investment into more comprehensive HIV screening and control initiatives in Madagascar.

## INTRODUCTION

Since the year 2000, UNAIDS data have demonstrated a rapid and exponential rise in HIV cases and AIDS-related deaths in Madagascar. Over the past 23 years, the number of people living with HIV in Madagascar has increased by over 4900%, the yearly incidence of new HIV cases has increased by approximately 1700%, and annual deaths due to AIDS have risen by greater than 2500% [1]. These rates appear to be accelerating, despite successful programs to combat HIV/AIDS in much of the rest of the African continent over the past two decades [2].

A major impediment to HIV control in Madagascar was a 2008 coup d’état that led to the suspension of many national health and foreign aid programs. This coup was followed by an interim government with high levels of instability, lacking sufficient resources for HIV research, prevention, and treatment [3]. As the Malagasy government recently began to reorganize and reinitiate public health programs, the COVID pandemic again suspended many initiatives, and SARS-CoV-2 became the central public health focus since its emergence [4].

Though the exponential increase in HIV rates is startling, HIV research in Madagascar is extremely limited, and available rates are largely based on what little data could be collected prior to the turmoil of recent decades. A 2020 meta-analysis on the current state of HIV research in Madagascar concluded that it is not possible to accurately determine the rate of HIV in the country based on the scarcity of data, and that the outputs of disease modeling should be interpreted with a high level of caution due to the lack of accurate epidemiologic data to use as inputs. This conclusion supports a growing consensus that official rates, based on spectrum modeling, could deeply underestimate the extent of HIV infection in Madagascar despite the rapid rise in cases that they already demonstrate [5]. A weakness of available studies is that most HIV data comes from select populations in urban centers, despite Madagascar having a majority rural population [5, 6]. Furthermore, health policies in Madagascar require HIV tests and antiretroviral medications to be supplied through the government [7]. Thus, clinics outside of well-connected urban centers or independent of the government are often unable to test patients for HIV or provide treatment when patients test positive [8].

Anthropologic studies have reported high rates of transactional sex across multiple regions of Madagascar, with approximately one-third of young females living in urban regions and two-thirds of those living in rural regions divulging engaging in relations for monetary compensation [9]. Relatedly, Madagascar carries one of the highest per capita burdens of gonorrhea, chlamydia, and syphilis infection the world [6,10], yet spectrum-model derived figures used by the Ministry of Health place the HIV prevalence uniquely low at 0.3% [5,11]. In addition to high sexually transmitted infection (STI) rates, another factor raising suspicion of elevated HIV rates is the extensive proportion of the populace involved in mining (>10%), where travel in pursuit of gem or precious metal deposits serves as a means of geographically disseminating HIV [12, 13].

The Institute Pasteur provided one major rural HIV prevalence estimate during a cross-sectional study they conducted in 2002 which reported an HIV rate of approximately 1% [6]. However, no large general-population screening studies have been published in the intervening two decades and there is considerable concern that rates could have increased during this time-interval beyond those observed by the Institute Pasteur.

Mada Clinics is a Malagasy government-certified healthcare center that provides free care for a large swath of rural Northern Madagascar, including nearly 75 towns, villages, and cities. Over the past seven years, the center noted an increasing burden of patients with fungal infections, recurrent fevers, and chronic weight loss, suggesting a clinical need to initiate HIV screening.

After completing a four-year process of certification with the Malagasy Ministry of Health, Mada Clinics initiated a screening and treatment protocol for HIV into all primary care visits in order to assist the Ministry of Health in collecting much needed population screening data. We posited that this study might yield valuable, general population, in-person testing data for models employed by the Ministry of Health. Furthermore, we aimed to assess how HIV rates may have changed in rural Madagascar over the past twenty years.

Given that Mada Clinics patient population includes adolescents and adults presenting for low acuity care, we posited that this group may serve as a more representative population than patients seen in private clinics or secondary/tertiary care centers. Building upon the work of the Institut-Pasteur, whose notable study surveyed 16 rural villages, this investigation including over one thousand patients from 73 towns, villages, and cities across Northern Madagascar. To our knowledge, this is the largest HIV prevalence study conducted among the general population of Madagascar to date.

## METHODS

### Patient enrollment

This study was conducted within a two-month interval between October 2022 and March 2023, during which all patients presenting to the clinic for routine care were asked whether they would like HIV screening. Patients were enrolled into the study daily until over 1,000 total patients were accrued. All participants were consented verbally prior to study enrollment, including consent from parents for minors. Prior to testing, all patients were counselled about basic HIV biology, its mechanism of transmission, clinical consequences of infection, and treatment options. Biographic and demographic data was collected from each subject and tabulated in an anonymous fashion. This study was conducted in accordance with the Declaration of Helsinki and was authorized by the Malagasy Ministry of Health in Antsiranana including approval by their Ethics Committee.

### Testing procedure

Patients who consented to study enrollment were tested using a two-tiered testing algorithm employing oral fluid or fingerstick blood samples. Screening was performed by using one of four commercially-available point-of-care assays for rapid detection of HIV-1/2 antibodies (**Supplemental Table 1**). Patients who screened positive for HIV underwent confirmatory testing using the SURE CHECK® HIV assay (Chembio Diagnostics, Medford, NY), an FDA-cleared device that provides superior sensitivity and specificity for HIV detection [14]. Additional confirmatory testing was typically conducted by the Ministry of Health leading positive patients to receive an average of three tests. All samples were obtained and tested by a trained medical staff member. Results were tabulated as positive or negative based on manufacturer instructions for each assay. Test kits were provided for use by the Malagasy Ministry of Health, with financial support from Mada Clinics when necessary.

**Supplemental Table 1.** Rapid screening assays for HIV.

| **Name** | **Manufacturer** | **Sample type (collection)** | **Methodology** |
| --- | --- | --- | --- |
| OraQuick® | OraSure Technologies | Oral fluid | Immunoassay |
| HIV-1/2 Antibody Rapid Test | Hightop Biotech | Whole blood (fingerstick) | Immunoassay |
| Diagnos HIV BI-DOT | J. Mitra & Co. | Whole blood (fingerstick) | Immunoassay |
| HIV 1.2.O Rapid Test Device | Rapid Labs Ltd. | Whole blood (fingerstick) | Immunoassay |

### Treatment and follow-up

In accordance with local health directives, patients who were confirmed to be HIV positive were registered with the local Ministry of Health in Anstiranana. Following guidance from a ministry-authorized infectious disease physician, HIV-positive patients were prescribed one of two available combination antiretroviral regimens (**Supplemental Table 2**). All medications were furnished from the Malagasy Ministry of Health and were dispensed by credentialed clinic staff. HIV-positive patients were scheduled for regular monthly follow-up at Mada Clinics. Patients were provided the option of referral to a subspecialty care center in Diego, where additional laboratory monitoring capabilities were available.

**Supplemental Table 2.** HIV treatment regimens.

| **Medications** |
| --- |
| <u>Regimen 1</u><br>Efavirenz 600mg<br>Lamivudine 300mg<br>Tenofovir disoproxil fumarate 300 mg |
| <u>Regimen 2</u><br>Dolutegravir 50mg<br>Lamivudine 300mg<br>Tenofovir disoproxil fumarate 300 mg |

## RESULTS

### Patient demographics and HIV testing results

1,026 patients presented for care during the study period, 1,019 of whom consented to undergo HIV screening and study enrollment. In total, 30 patients tested positive on all screening and confirmatory tests, representing an overall HIV prevalence rate of 2.94% (**Table 1**). 66.7% (20/30) of positive cases occurred in females, representing a 3.47% positivity rate in female patients and a 2.26% positivity rate in male patients (**Figure 1A**). HIV positivity was greatest in patients in the 20s to 30s age range, with a mean age of 35.2 years (**Figure 1B**). Of note, only one individual under the age of 18 tested positive for HIV.

**Table 1.** Patient characteristics and overall HIV prevalence.

| Age |  |
| --- | --- |
| N | 1019 |
| Mean (SD) | 36.7 (15.2) |
| Median | 34 |
| Range | 13, 82 |
| Sex, n (%) |  |
| Female | 577 (56.6%) |
| Male | 442 (43.4%) |
| Prevalence |  |
| HIV positive | 30 (2.9%) |
| HIV negative | 989 (97.1%) |

**Figure 1:**
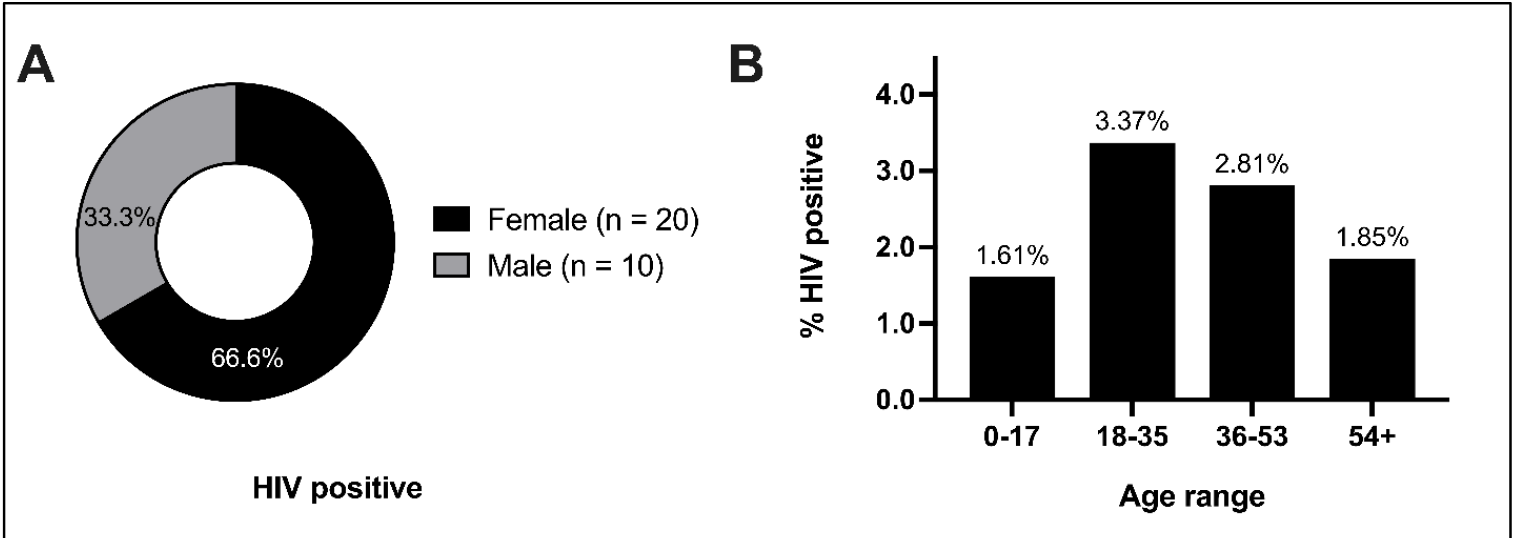
Demographics of HIV positive individuals. **A)** Total number of HIV-positive individuals enumerated by sex, and **B**) rate of HIV positivity in study population by age group.

### Geographic and sociographic patterns

**Supplemental Figure 1:**
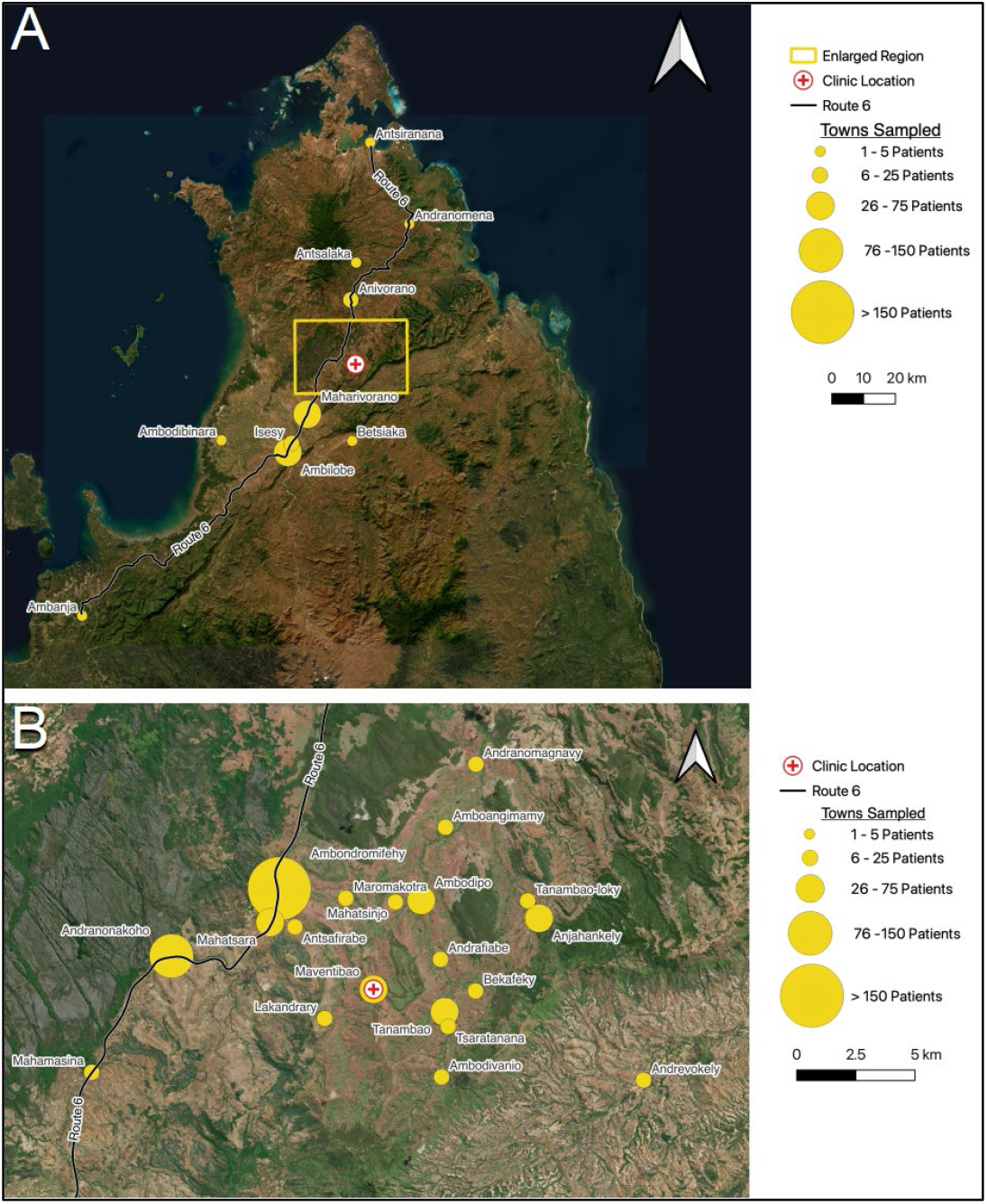
Geographic distribution of HIV prevalence. **A)** Large-scale, and **B**) detailed view of villages, towns, and cities from which HIV-positive individuals were identified. Photos courtesy of Google Earth.

Patients presented from 73 different towns, villages, or cities. While the majority of patients lived in rural villages, 8.2% (84) of patients presented from cities or suburbs (“urban”) (**Figure 2A**). Among this urban population, the HIV prevalence was significantly greater, **13.1**% than among the rural population, where 2.03% were found to be HIV-positive. We hypothesized that increased mobility and access to major routes of transportation could be associated with HIV infection. One major transportation route is Route Nationale 6 (RN6), the primary highway in Northern Madagascar that spans 706 kilometers from Antsiranana to Ambondromamy. The majority of patients were found to live along RN6, and among this population, the HIV prevalence was significantly greater compared to those living away from RN6 (**Figure 2B**). Strikingly, all (10/10) HIV-positive men lived along RN6, with an overall HIV rate of 3.40% in this segment. In contrast, no males living away from RN6 (0/148) tested positive for HIV, despite this demographic constituting a major subpopulation of the study.

**Figure 2:**
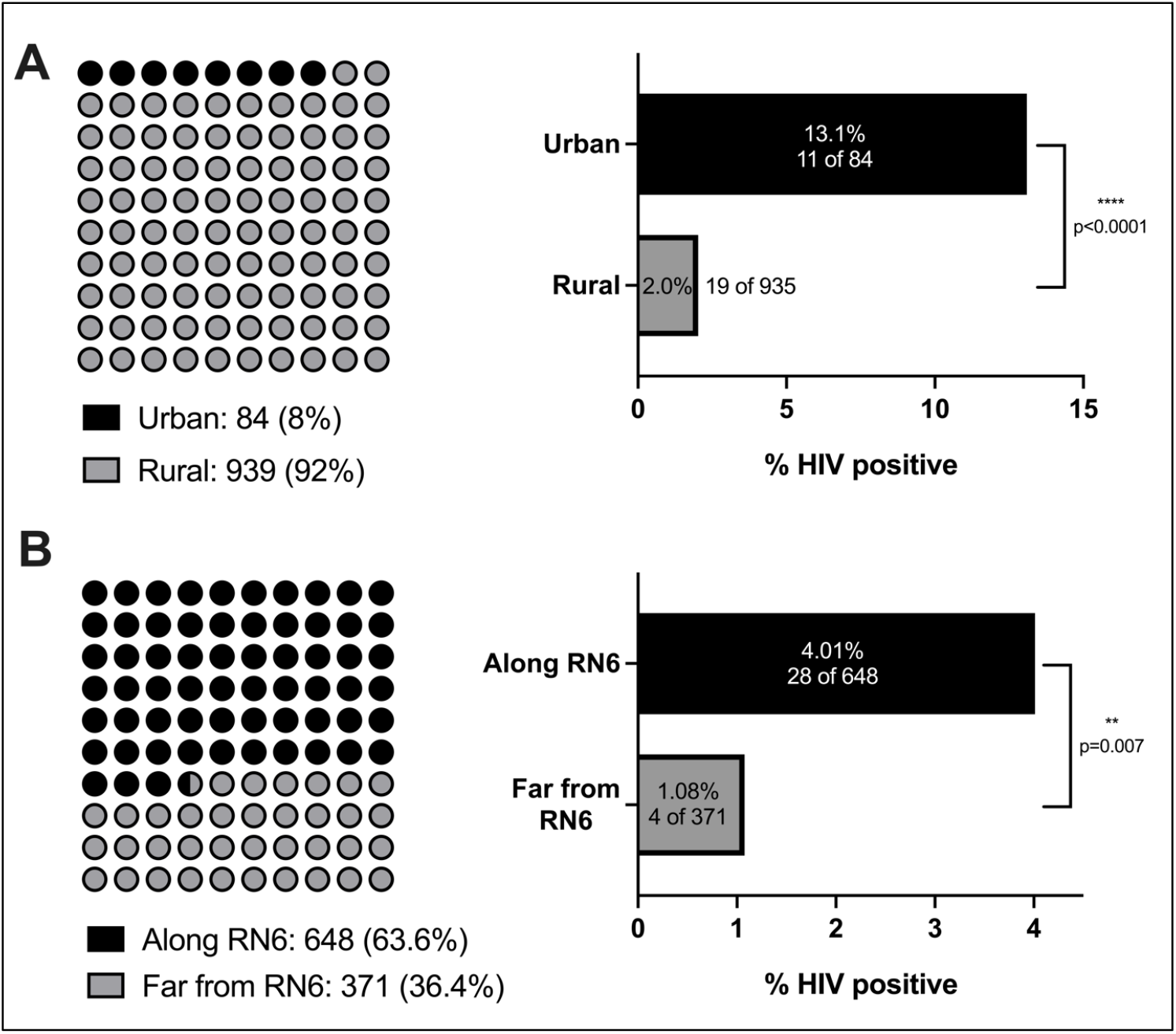
Geographic distribution of HIV prevalence. **A)** Total number of study participants from urban or suburban locales (black) versus rural villages (grey) and proportion of HIV-positive individuals originating from each geography. **B)** Number of individuals living close to (black) or far from RN6 (grey) and percent of HIV-positive individuals originating from each locale. Significance was determined using two-tailed Fisher’s exact test (defined as p<0.05).

An additional factor we hypothesized to be associated with HIV status was occupation. Participants in the study practiced 30 unique professions. Patients who tested positive for HIV were found among eight total occupations, of which agriculture and mining encompassed the greatest number of HIV-positive individuals, respectively (**Figure 3A**). Relatively, HIV prevalence was highest (66.67%) among sex workers, though the sample size of this group was limited (**Figure 3B**). Although both agriculture and mining are major common occupations in Madagascar, there was a significantly increased relative-risk of HIV infection in miners (**Figure 3C**). Indeed, when segmented by gender, the majority of HIV-positive men practiced mining (7/10), while no male farmers were HIV-positive (**Figure 4**). In contrast, there was a greater variety of occupations among HIV-positive females.

**Figure 3:**
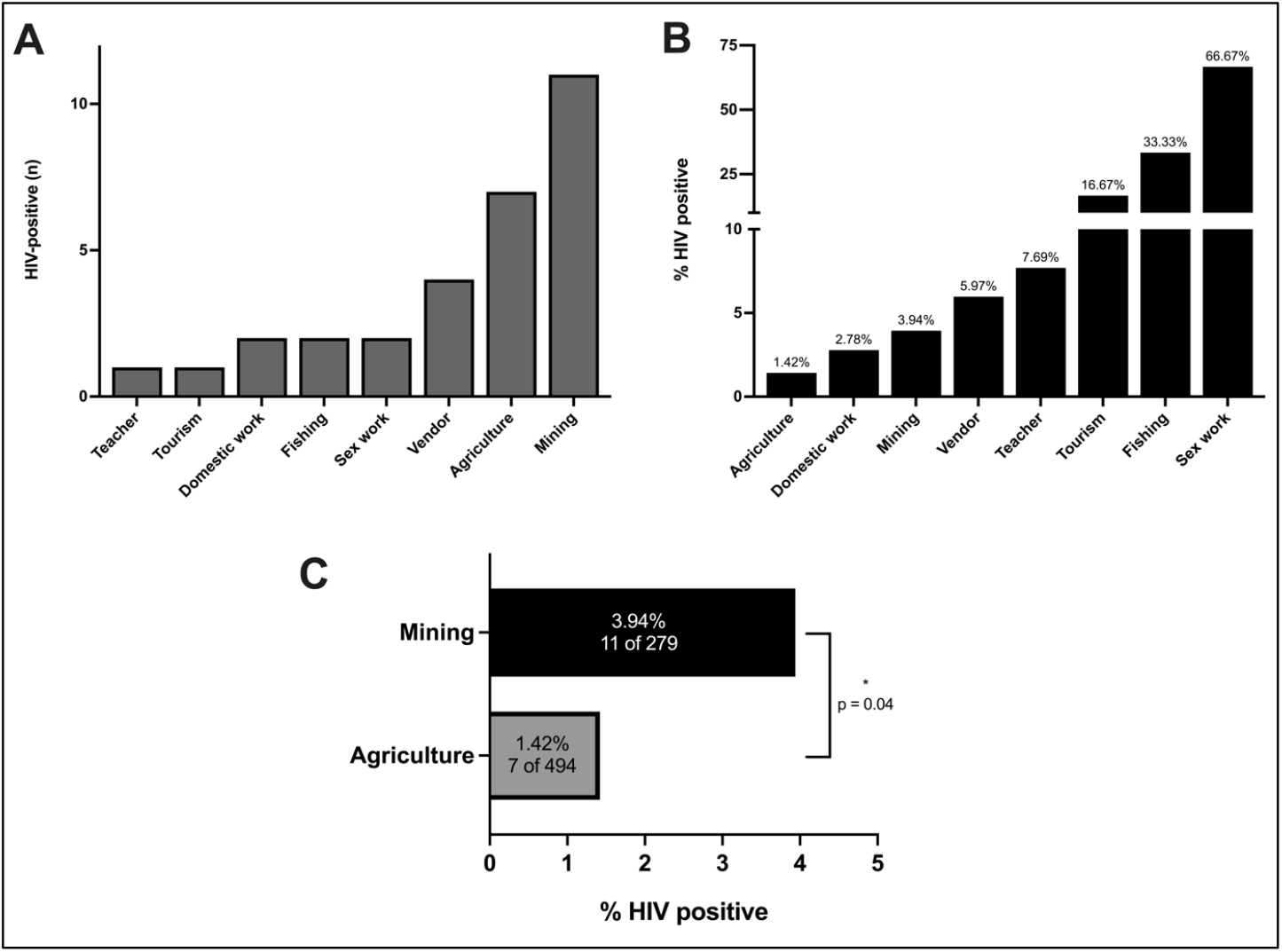
HIV status by occupation. **sA)** Occupations held by individuals who tested positive for HIV, **B)** percent HIV-positive individuals by occupation, and **C)** HIV-positive individuals in mining versus agriculture. Significance was determined using two-tailed Fisher’s exact test (defined as p<0.05).

**Figure 4:**
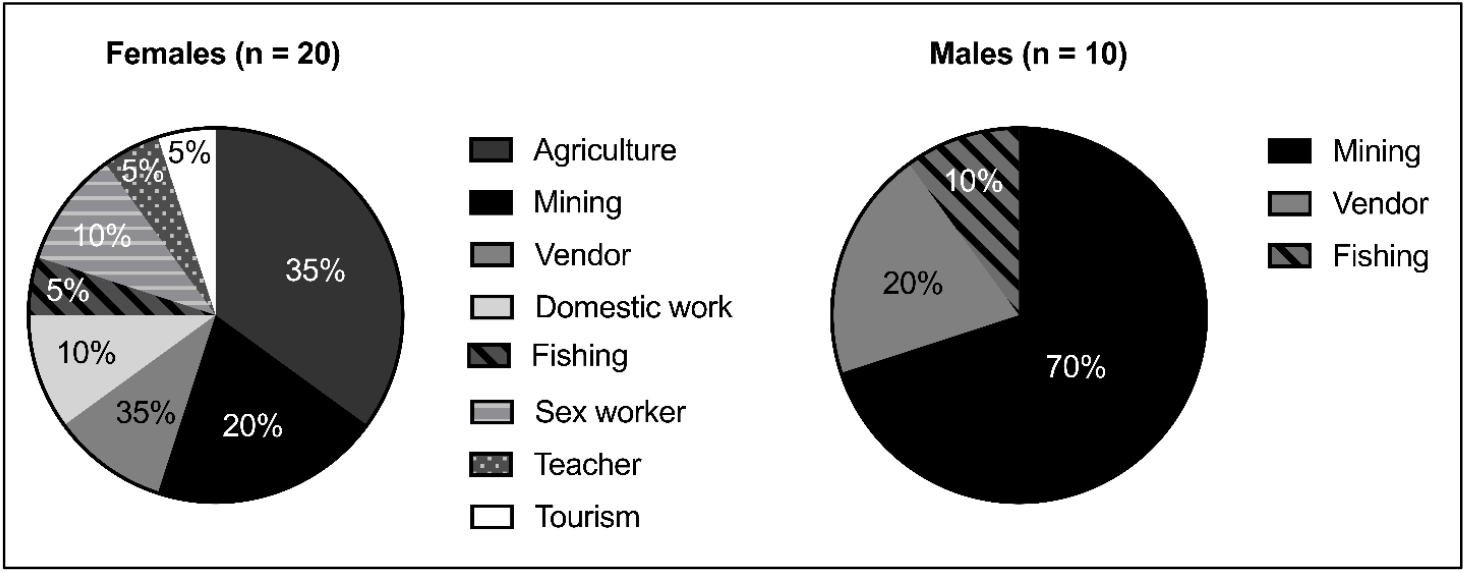
Occupations of HIV-positive individuals divided by gender.

### Presenting context of HIV-positive individuals

Patients enrolled in the study presented for care due to a variety of clinical concerns. The most common presenting chief complaints among HIV-positive patients were another STI (n=8), skin infection (n=7), constitutional symptoms (n=3), and contraception (n=2), respectively. Of these, constitutional symptoms (which included chronic fatigue and weight loss) demonstrated the highest HIV positivity rate of these conditions, with 60% (3/5) of these patients testing positive. Comparatively, 20% (2/10) of those presenting for a contraceptive visit tested positive for HIV (2/10). Of note, no pregnant female patients tested positive for HIV (0/76), suggesting that HIV screening in the prenatal setting alone may not serve as a representative screening strategy for the total population.

## DISCUSSION

To our knowledge, this study represents the largest general population HIV prevalence study conducted in Madagascar to date. The overall prevalence rate observed, 2.94%, is substantially higher than the spectrum-model generated rate of 0.3% used by the Ministry of Health, which may have been biased by the scarcity of primary patient test data. Our results largely agree with findings reported by the Institut Pasteur in 2002[6]. Although we observed a higher HIV prevalence of 2.9% as compared to 1%, this increase over a 21-year span could be expected given the low awareness of HIV status among HIV-positive individuals (15%), and the low rates of HIV-positive individuals who are on active antiretroviral therapy (15%) reported in UNAIDS data [1]. In the largely rural population surveyed in this study, 0% of HIV-positive individuals knew their status prior to this study, and 0% of HIV-positive individuals were on antiretroviral treatment.

Geography was a key factor we observed to influence HIV prevalence. In comparison to the Institut Pasteur’s study, which focused exclusively on rural individuals, our study included patients from rural villages as well as urban and suburban locales. While the HIV rate we observed in rural patients (2.0%) was closer the rate observed by the Institut Pasteur in 2002, we noted a six-fold higher rate in urban patients (13.1%) than rural patients, and their inclusion in this study likely accounts for a portion of the elevation in rates observed. We also report a significant association between HIV status and proximity to RN6, the major highway that serves northern Madagascar. Given the evidence of an elevated HIV prevalence in urban centers, we hypothesize that such a thoroughfare might provide a means of HIV dissemination, particularly for individuals who often travel between different towns and cities.

HIV prevalence also varied significantly by occupation, with professional sex work correlating most strongly with HIV-positivity. Interestingly though, careers such as shopkeeping and tourism also demonstrated a high HIV burden. One link between these occupations and potential HIV positive status is the need for these individuals to frequently travel to urban centers, whether to restock goods or provide services. In absolute terms, mining had the greatest number of HIV-positive individuals. While this is not surprising given mining is a major industry in Madagascar, this also provides credence to the importance of an individual’s mobility on HIV risk. Miners in Madagascar often move residence based on which locale provides the greatest economic opportunity at a given time [15]. Further work is warranted to investigate whether HIV transmission also increases during mining “boom” cycles.

Adding further evidence to these observations, we found that isolation from RN6 and practicing agriculture were negatively associated with HIV positivity. In fact, no males living in villages away from RN6 and no males who primarily practiced agriculture were found to be HIV-positive in our study, despite large sample sizes. This association was also seen among females, though to a lesser degree. Overall miners were 2.8 times as likely to test positive for HIV as compared to those with the more stationary occupation of farming.

Overall women were 1.5 times as likely as men to test positive for HIV in our study. A contributing explanation is likely the higher physiologic risk of infection faced by women due to increased membrane surface area exposure to infection [16]. Additionally, whereas 70% of HIV positive males practiced mining, HIV positive females practiced a much greater variety of occupations than men **(figure 4)**. A potential factor linking the increased risk of infection faced by women and the greater variety of occupations observed could be the high rates of transactional sex that are especially common and often normalized in rural Madagascar, which often supplement incomes for young females [15]. Thus, this additional risk factor could elevate HIV risk regardless of primary occupation. Adding evidence to this, we observed a 66% higher risk of HIV infection in women under the age of 35 as compared to those above this age range and a 3.58 times higher rate of HIV infection in unmarried women as compared to those who were married. These trends were not similar for men, who conversely demonstrated a higher risk of HIV infection with increased age.

There are a few key limitations to our study: Although we sampled a geographically varied population across northern Madagascar, our study did not garner individuals from central or southern Madagascar. Because many cultural and economic distinctions may exist between these regions, the prevalence and rate of HIV transmission may also differ. Furthermore, our testing focused primarily on patients who presented for free, routine primary care. Future efforts might employ screening strategies that are executed directly in the community to further decrease the risk of any sampling bias.

Nevertheless, our findings mark a major advance in the understanding of the extent of HIV across Madagascar. Despite limited HIV data to date, investigators have increasingly begun to question whether a generalized HIV epidemic is on the horizon [5]. Our findings, capturing not only a largely rural population but also individuals from urban centers, strongly suggest that an HIV epidemic is already present within Northern Madagascar, and may be present across the country as a whole. During the process of completing this study, we received numerous requests from other healthcare centers, including government infectious disease clinics, for assistance in providing HIV testing kits. This suggests healthcare centers in this region have minimal testing capacity for HIV, further compounding the need for greater investment into testing infrastructure and treatment programs. Based on our findings, we strongly advise the international community to support Madagascar in strengthening HIV testing and treatment programs nationwide. The burden of illness we observed both at Mada Clinics and at hospitals in Antsiranana during this study, leads us to conclude that many people are likely dying each day from HIV/AIDS in Madagascar without notice or recognition. The success of programs to combat HIV/AIDS across the African continent in recent decades clearly demonstrates that such tragedies are avoidable [2].

## Data Availability

All data produced in the present study are available upon reasonable request to the authors.

